# Divergent Trends in Stroke and Ischemic Heart Disease Mortality and Disability in Western Sub-Saharan Africa Compared with Global Progress, 1990–2023

**DOI:** 10.64898/2026.08.19.26360860

**Authors:** Prince Ankrah-Twumasi, Jeffrey Jerry Vladimir Ofori, Prince Pekyi-Boateng, Yaw Twerefour, Dorcas Sackey

**Affiliations:** School of Medicine, Medical Sciences and Nutrition, University of Aberdeen, Aberdeen, United Kingdom; Department of Public Health, Tema General Hospital, Tema, Ghana; Department of Neurology, University Of Utah Health, SLC, UT, USA; Ghana Health Service, Accra, Ghana; Harvard Medical School, Boston, MA, USA

**Keywords:** Stroke, Ischemic heart disease, Cardiovascular disease, Western Sub-Saharan Africa, DALYs, Global Burden of Disease, SDI

## Abstract

**Background:** Cardiovascular disease remains the leading cause of death worldwide, yet progress in reducing its burden has not been shared equally across regions. Sub-Saharan Africa has previously been identified as the only world region where age-standardized cardiovascular mortality failed to decline, but long-term, disease-specific trends in Western Sub-Saharan Africa (WSSA) remain poorly characterized.

**Methods:** We conducted an ecological trend analysis using Global Burden of Disease (GBD) 2023 data to evaluate age-standardized mortality and disability-adjusted life years (DALYs) for stroke and ischemic heart disease (IHD) in WSSA and globally from 1990 to 2023. Linear and segmented regression assessed long-term trends and breakpoints, risk factor attribution examined six major cardiovascular risk factors, and Pearson correlation evaluated associations between the Socio-demographic Index (SDI) and mortality.

**Results:** Global stroke and IHD mortality declined by 51.7% and 38.2%, respectively, between 1990 and 2023. In WSSA, stroke mortality declined by only 21.8%, while IHD mortality increased by 3.3%. Segmented regression identified a breakpoint in IHD mortality around 2007, after which the trend reversed from declining to increasing. High systolic blood pressure was the leading attributable risk factor for both diseases, while obesity, ambient air pollution, and elevated fasting glucose showed the largest relative increases. SDI rose 69.5% in WSSA but correlated strongly only with stroke mortality (r = −0.87), not IHD (r = 0.21).

**Conclusions:** WSSA is falling behind global cardiovascular progress, with IHD mortality reversing course despite substantial socioeconomic development. Targeted investment in hypertension control, cardiometabolic risk reduction, and cardiovascular care capacity is urgently needed to prevent this divergence from deepening.

## 1. Introduction

Cardiovascular disease remains the leading cause of death worldwide, and stroke and ischemic heart disease (IHD) together account for the majority of this burden. The most recent Global Burden of Disease (GBD) 2021 estimates report that stroke alone caused 7.3 million deaths in 2021 (10.7% of deaths worldwide), making it the third leading cause of death globally after IHD and COVID-19, with 87.2% of stroke deaths and 89.4% of stroke-related disability-adjusted life years (DALYs) occurring in low-and middle-income countries.¹ IHD accounted for a further 9.0 million deaths and 188.4 million DALYs in the same year.² Although global age-standardized cardiovascular mortality has declined over the past three decades, this progress has not been shared equally across regions: earlier Global Burden of Disease estimates identified Sub-Saharan Africa as the only world region where age-standardized cardiovascular mortality failed to decline between 1990 and 2013,³ and more recent analyses confirm that the absolute number of cardiovascular deaths in the region has continued to rise even as global totals fall.⁴

Stroke and IHD, while sharing many of the same modifiable risk factors, follow distinct epidemiological trajectories with different implications for prevention and care. Stroke has historically dominated the cardiovascular disease burden in Sub-Saharan Africa, with substantially more deaths attributed to stroke than to IHD as recently as the early 2010s.³ This balance is now shifting: Taha and colleagues described ischemic heart disease in Africa as undergoing an “overnight epidemiological transition,” finding that IHD has already become the leading cause of death among men, and the second leading cause among women, over 60 years of age in the African region.⁵ Because the two conditions differ in risk-factor sensitivity, case fatality, and the health-system resources required for effective prevention and treatment, tracking them separately, rather than as a single combined cardiovascular category, is essential for understanding how the region’s disease burden is evolving.

This shifting balance reflects a broader epidemiological transition reshaping cardiovascular disease across Sub-Saharan Africa. Using GBD 2019 data, Xia and colleagues found that although the global age-standardized incidence rate of IHD declined between 1990 and 2019, it rose in Western Sub-Saharan Africa specifically, a pattern the authors linked to the region’s rapid socioeconomic transition.⁶ This finding is consistent with classical models of epidemiological transition, in which stroke mortality tends to decline relatively early in the course of socioeconomic development, driven largely by improved blood pressure control, while IHD mortality rises later as populations adopt more westernized diets and increasingly sedentary lifestyles.⁷ Minja and colleagues likewise concluded that non-communicable diseases, including the atherosclerotic conditions that underlie IHD, are on track to overtake communicable diseases as the leading cause of death across the African continent within the current decade.⁸

Despite this evidence of an emerging cardiovascular transition, long-term, region-specific data describing how stroke and IHD burden have actually evolved in Western Sub-Saharan Africa (WSSA) remain limited. A 2026 assessment of healthcare system readiness across Sub-Saharan Africa found that systematic, country-level evaluations pairing IHD burden with healthcare capacity remain scarce, leaving substantial gaps for evidence-based policy.⁹ This gap is compounded by longstanding methodological challenges in estimating cardiovascular disease burden in the region: because GBD estimates rely heavily on modeled rather than directly observed data, their accuracy depends on the quality and completeness of underlying vital registration, which remains sparse across many WSSA countries.¹⁰ Without dedicated, long-term trend analyses specific to Western Sub-Saharan Africa, it remains unclear whether the region is following, lagging behind, or diverging from global progress in reducing stroke and IHD mortality and disability, information that is essential for setting regional prevention priorities and strengthening data-driven health policy.⁸

To address this gap, we conducted an ecological trend analysis using data from the Global Burden of Disease (GBD) 2023 study to characterize long-term trends in age-standardized mortality and disability-adjusted life years attributable to stroke and ischemic heart disease in Western Sub-Saharan Africa between 1990 and 2023, and to compare these trends with corresponding global estimates. We further examined trends in major attributable cardiovascular risk factors and the relationship between socio-demographic development and disease-specific mortality, with the goal of informing region-specific cardiovascular prevention strategies.

## 2. Methods

### Study Design

This ecological trend analysis used publicly available data from the Global Burden of Disease (GBD) 2023 study to evaluate long-term trends in age-standardized mortality and disability attributable to stroke and ischemic heart disease (IHD) in Western Sub-Saharan Africa (WSSA) between 1990 and 2023. Trends observed in WSSA were compared with corresponding global estimates to assess regional progress relative to worldwide patterns.

### Study Setting

Western Sub-Saharan Africa comprises 16 countries: Benin, Burkina Faso, Cabo Verde, Côte d’Ivoire, The Gambia, Ghana, Guinea, Guinea-Bissau, Liberia, Mali, Mauritania, Niger, Nigeria, Senegal, Sierra Leone, and Togo. Regional estimates provided by the GBD study were used for all WSSA analyses.

### Data Source

Data were obtained from the GBD 2023 study, coordinated by the Institute for Health Metrics and Evaluation. The GBD provides standardized estimates of mortality, disability, risk factor attribution, and socio-demographic development across countries and regions using a comprehensive framework that synthesizes data from vital registration systems, censuses, surveys, disease registries, and epidemiological studies.

The following GBD-derived datasets were analyzed:

1. Age-standardized mortality rates.
2. Age-standardized disability-adjusted life year (DALY) rates.
3. Risk factor–attributable DALY rates.
4. Socio-demographic Index (SDI) estimates.

All rates were reported per 100,000 population.

### Outcomes of Interest

The primary outcome was age-standardized mortality attributable to stroke and ischemic heart disease. Age-standardized mortality rates were obtained directly from the Global Burden of Disease (GBD) 2023 study and are reported per 100,000 population. Age standardization facilitates comparisons across populations and over time by accounting for differences in age structure.

Secondary outcomes included age-standardized DALY rates, which represent the combined burden of years of life lost due to premature death and years lived with disability.

### Risk Factor Attribution Analysis

To explore potential drivers of observed trends, DALY rates attributable to six established cardiovascular risk factors were examined:

- High systolic blood pressure
- Dietary risks
- High body-mass index
- High fasting plasma glucose
- Ambient particulate matter pollution
- Tobacco use

For each disease, annual attributable DALY rates were extracted for 1990–2023. Absolute rates in 2023 and percentage changes between 1990 and 2023 were calculated to assess temporal changes in risk factor burden.

### Socio-demographic Development Analysis

Socio-demographic Index (SDI) estimates were extracted for both WSSA and the global population. SDI is a composite indicator of development incorporating income per capita, educational attainment, and fertility among individuals younger than 25 years.

Temporal changes in SDI between 1990 and 2023 were calculated. Pearson correlation analyses were subsequently performed to evaluate associations between annual SDI values and age-standardized mortality rates for stroke and ischemic heart disease in WSSA.

### Statistical Analysis

Descriptive analyses were performed to summarize trends in mortality, DALYs, SDI, and risk factor attribution from 1990 to 2023.

Percentage change between 1990 and 2023 was calculated as:

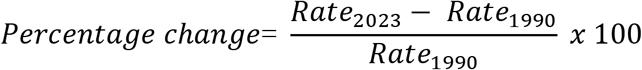

Linear regression models were fitted to quantify long-term temporal trends in age-standardized mortality rates for stroke and ischemic heart disease. Age-standardized mortality rate was modeled as a function of calendar year. Regression coefficients (β), coefficients of determination (R²), and p-values were reported. The regression coefficient (β) represents the average annual change in age-standardized mortality rate per 100,000 population.

To identify potential changes in trend trajectories over time, segmented regression analyses were conducted using the segmented package in R. Breakpoints were estimated using iterative procedures that identify statistically optimal change-points in temporal trends. Pre-breakpoint and post-breakpoint slopes were subsequently calculated to characterize changes in age-standardized mortality trajectories over time.

Pearson correlation coefficients (r) with corresponding 95% confidence intervals were used to assess associations between SDI and disease-specific mortality rates.

All analyses were conducted using R statistical software (version 4.5.1). Statistical significance was defined as a two-sided p-value < 0.05.

## 3. Results

### Trends in Age-Standardized Mortality in Western Sub-Saharan Africa and Globally

Between 1990 and 2023, age-standardized mortality rates for both stroke and ischemic heart disease (IHD) declined substantially at the global level. Global age-standardized stroke mortality decreased from 157.0 to 75.8 deaths per 100,000 population, representing a 51.7% reduction. Similarly, global age-standardized IHD mortality declined from 161.0 to 99.7 deaths per 100,000 population, corresponding to a 38.2% reduction.

In contrast, progress in Western Sub-Saharan Africa (WSSA) was markedly slower. Age-standardized stroke mortality declined from 147.0 to 115.0 deaths per 100,000 population, representing a 21.8% reduction over the study period. Age-standardized IHD mortality increased slightly from 93.7 to 96.8 deaths per 100,000 population, corresponding to a 3.3% increase between 1990 and 2023 (Table 1).

**Table 1.** Changes in Age-Standardized Mortality and Disability Burden from Stroke and Ischemic Heart Disease in Western Sub-Saharan Africa and Globally, 1990–2023. Outcome rates are expressed per 100,000 population.

| Outcome | Region | Disease | Rate in 1990 | Rate in 2023 | Percentage Change (%) |
| --- | --- | --- | --- | --- | --- |
| Mortality | Global | Stroke | 157 | 75.8 | -51.7 |
| Mortality | Western Sub-Saharan Africa | Stroke | 147 | 115 | -21.8 |
| Mortality | Global | Ischemic heart disease | 161 | 99.7 | -38.2 |
| Mortality | Western Sub-Saharan Africa | Ischemic heart disease | 93.7 | 96.8 | 3.3 |
| DALYs | Global | Stroke | 3356 | 1737 | -48.2 |
| DALYs | Western Sub-Saharan Africa | Stroke | 3447 | 2716 | -21.2 |
| DALYs | Global | Ischemic heart disease | 3277 | 2129 | -35 |
| DALYs | Western Sub-Saharan Africa | Ischemic heart disease | 1989 | 2039 | 2.5 |

**Abbreviations:** DALYs, disability-adjusted life years.

Visual inspection of mortality trajectories demonstrated sustained global declines for both diseases throughout the study period. In WSSA, stroke mortality declined at a slower pace, while IHD mortality exhibited a more complex pattern characterized by an initial decline followed by subsequent increases during the later years of observation (Figures 1 and 2).

**Figure 1.**
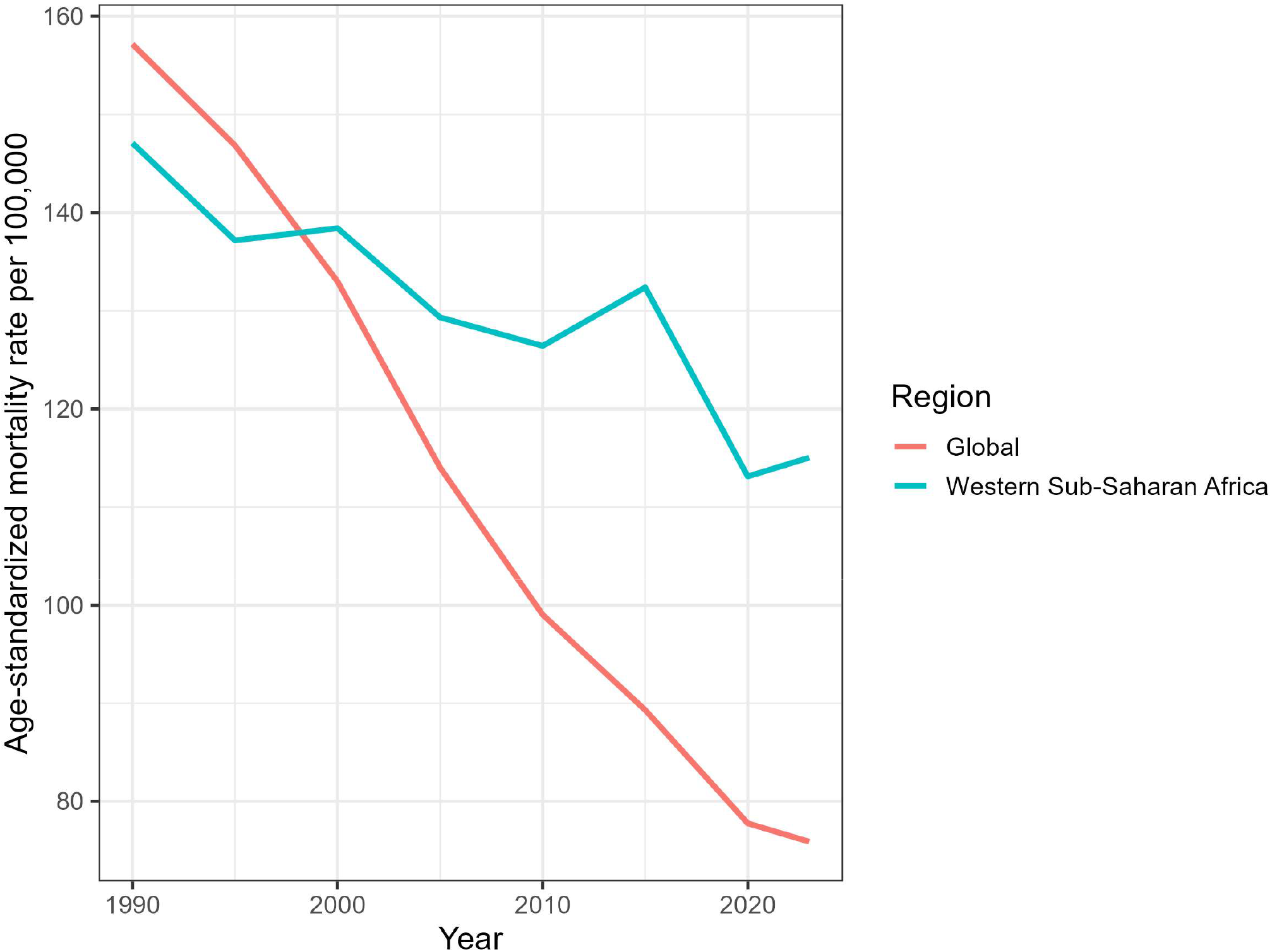
Trends in age-standardized stroke mortality rates in Western Sub-Saharan Africa and globally, 1990–2023. Mortality rates are expressed per 100,000 population and were obtained from the Global Burden of Disease 2023 study.

**Figure 2.**
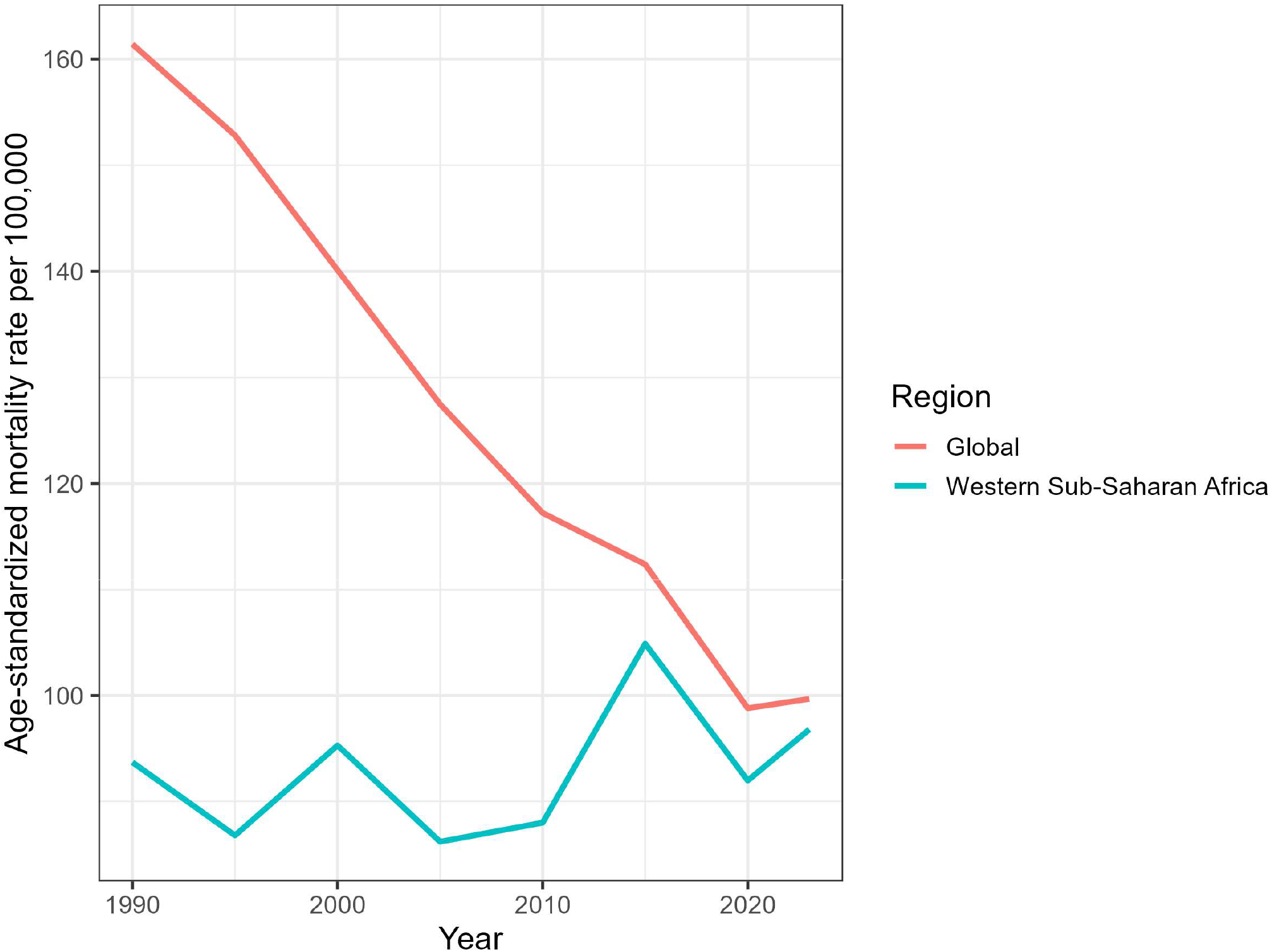
Trends in age-standardized ischemic heart disease mortality rates in Western Sub-Saharan Africa and globally, 1990–2023. Mortality rates are expressed per 100,000 population and were obtained from the Global Burden of Disease 2023 study.

### Linear Trend Analysis of Age-Standardized Mortality

Linear regression analyses confirmed significant downward trends in stroke mortality both globally and within WSSA. Globally, age-standardized stroke mortality declined by an average of 2.73 deaths per 100,000 population annually (β = −2.725, R² = 0.976, p < 0.001). In WSSA, stroke mortality also declined significantly, although at a substantially slower rate of 1.02 deaths per 100,000 population annually (β = −1.024, R² = 0.803, p < 0.001).

For IHD, global mortality demonstrated a strong and consistent decline over time (β = −1.866, R² = 0.965, p < 0.001). In contrast, no significant long-term linear trend was observed for age-standardized IHD mortality in WSSA (β = +0.083, R² = 0.019, p = 0.432), indicating an absence of sustained improvement during the study period (Table 2).

**Table 2.** Linear Regression Analysis of Long-Term Trends in Age-Standardized Mortality Rates, 1990–2023.

| Disease | Region | Regression Coefficient ($\beta$) | R<sup>2</sup> | p-value |
| --- | --- | --- | --- | --- |
| Stroke | Global | -2.725 | 0.976 | <0.001 |
| Stroke | Western Sub-Saharan Africa | -1.024 | 0.803 | <0.001 |
| Ischemic heart disease | Global | -1.866 | 0.965 | <0.001 |
| Ischemic heart disease | Western Sub-Saharan Africa | 0.083 | 0.019 | 0.432 |

### Temporal Changes in Mortality Trajectories

Segmented regression analyses identified significant changes in mortality trajectories within WSSA (Table 3).

**Table 3.**
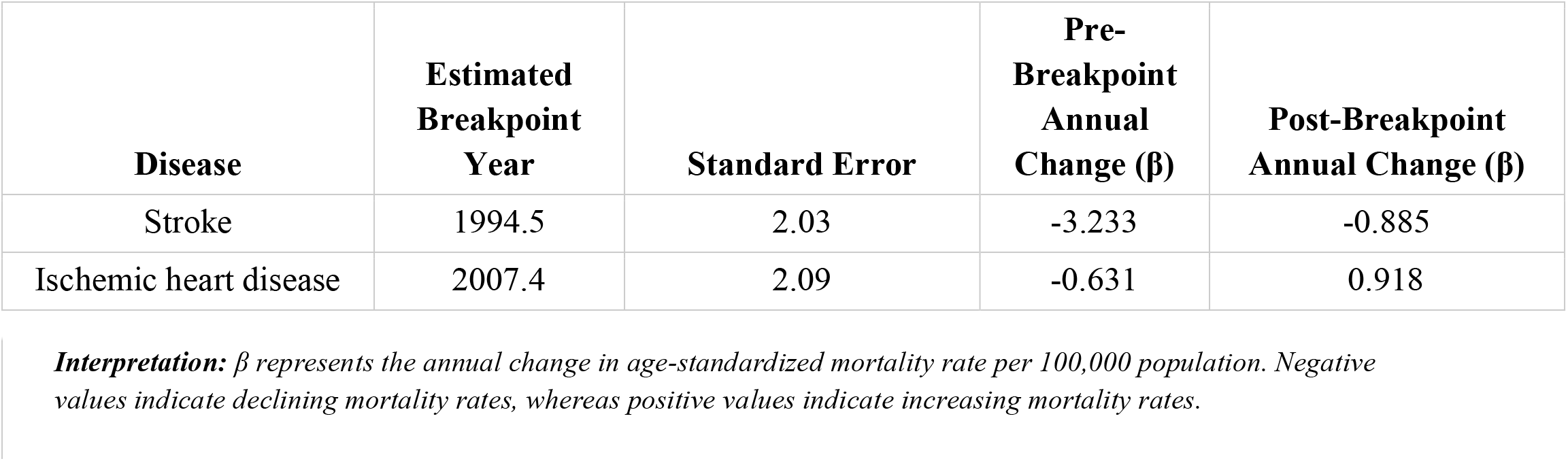
Segmented Regression Analysis of Mortality Trends in Western Sub-Saharan Africa, 1990–2023.

For stroke, a breakpoint was identified around 1995 (estimated breakpoint year 1994.5 ± 2.0 years). Prior to this breakpoint, age-standardized stroke mortality declined rapidly (β = −3.233 per year). After 1995, mortality continued to decline but at a substantially slower rate (β = −0.885 per year), indicating a marked attenuation in the pace of improvement (Figure 5).

For IHD, a breakpoint was identified around 2007 (estimated breakpoint year 2007.4 ± 2.1 years). Prior to 2007, age-standardized IHD mortality declined annually (β = −0.631 per year). Following this breakpoint, the direction of the trend reversed, with mortality increasing at approximately 0.918 deaths per 100,000 population annually. This finding suggests a reversal of earlier gains in age-standardized IHD mortality reduction within the region (Figure 6).

### Trends in Disability Burden

Patterns observed for disability-adjusted life years (DALYs) closely mirrored mortality trends.

Globally, age-standardized DALY rates declined from 3356 to 1737 per 100,000 population for stroke, representing a 48.2% reduction, and from 3277 to 2129 per 100,000 population for IHD, representing a 35.0% reduction.

In WSSA, stroke DALY rates declined from 3447 to 2716 per 100,000 population, corresponding to a 21.2% reduction. In contrast, IHD DALY rates increased from 1989 to 2039 per 100,000 population, representing a 2.5% increase over the study period.

The concordance between mortality and DALY trends indicates that the observed divergence between WSSA and global cardiovascular outcomes extended beyond mortality and was also reflected in disability burden (Table 1; Figures 3 and 4).

**Figure 3.**
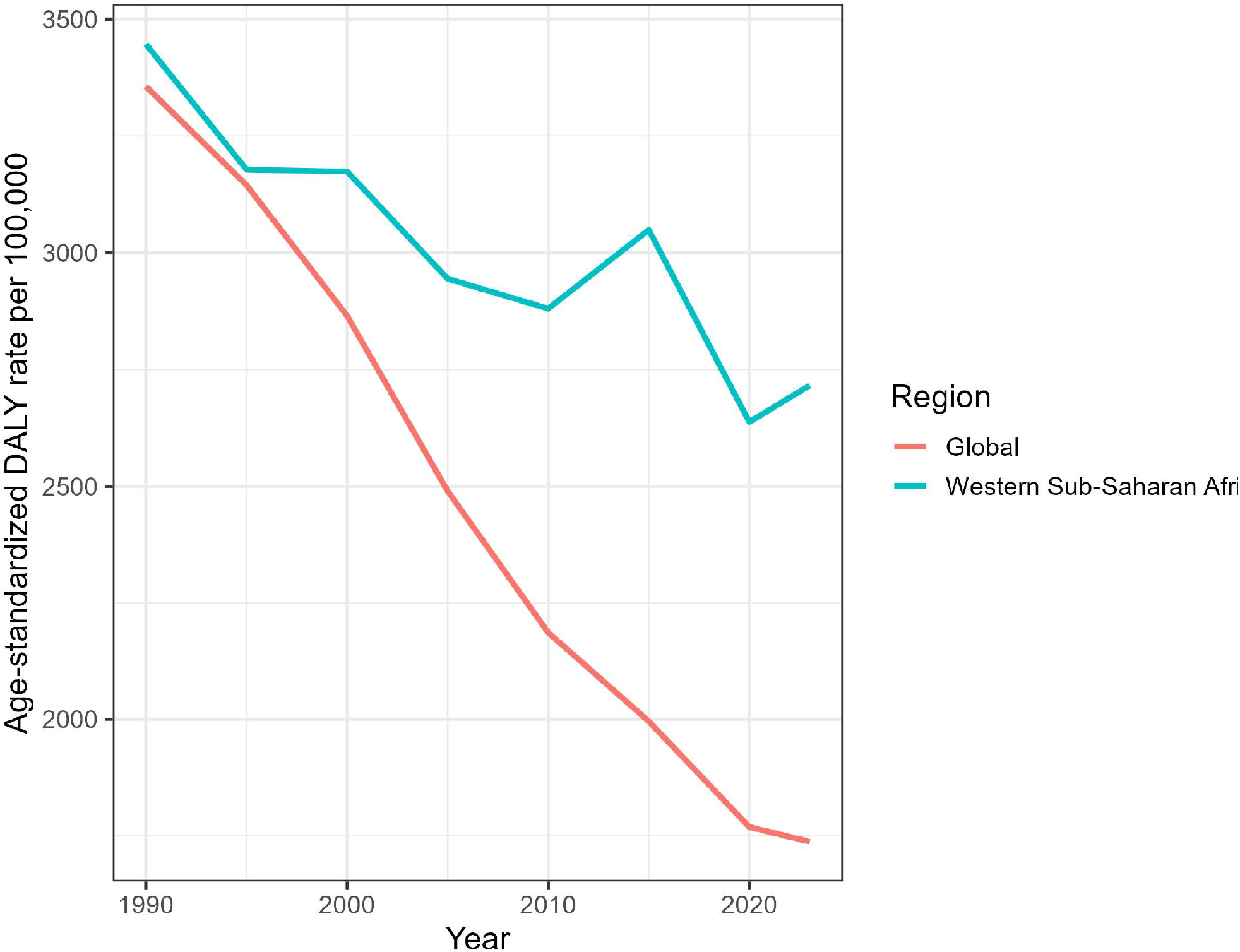
Trends in age-standardized stroke DALY rates in Western Sub-Saharan Africa and globally, 1990–2023. DALY rates are expressed per 100,000 population and were obtained from the Global Burden of Disease 2023 study.

**Figure 4.**
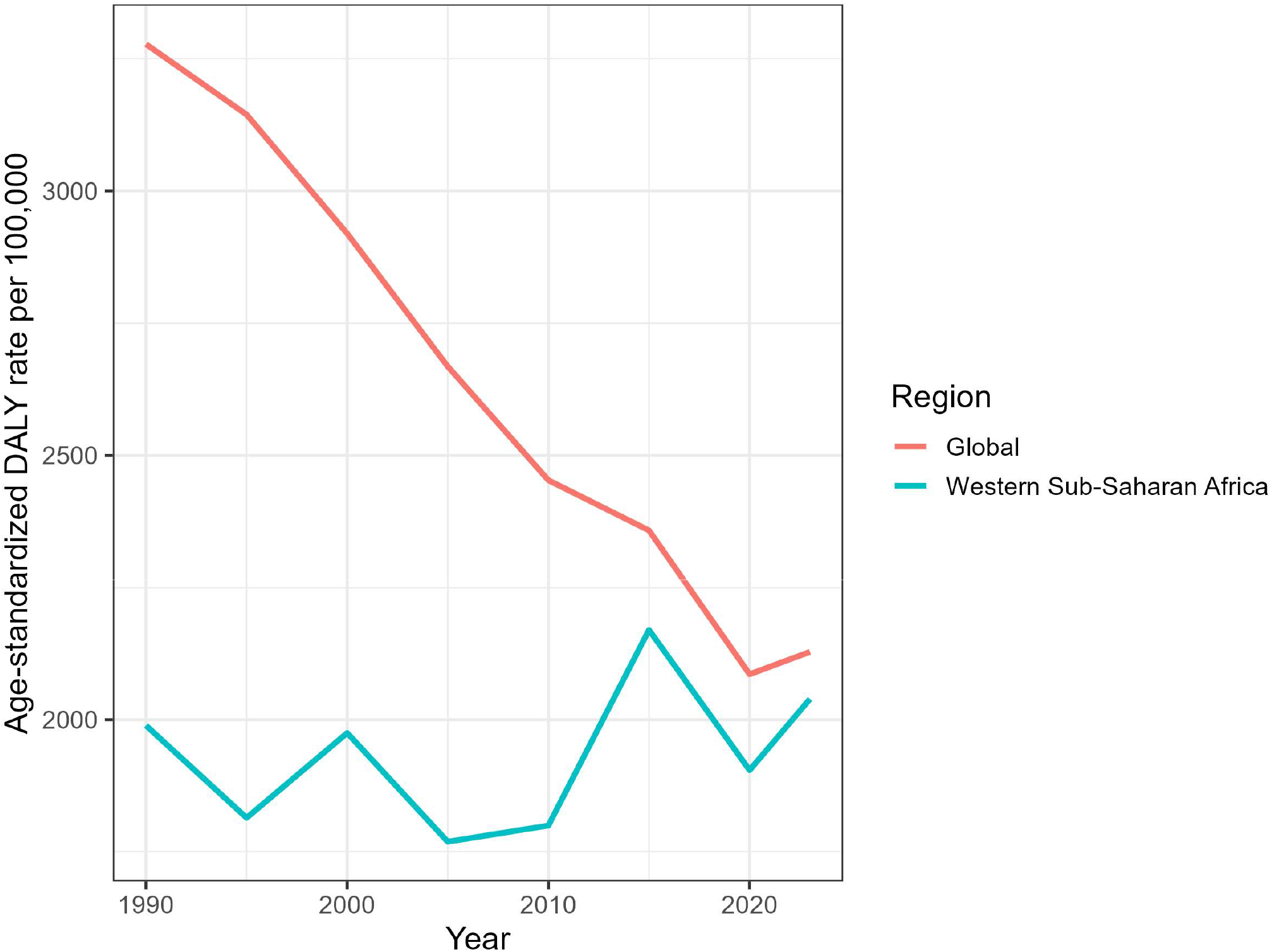
Trends in age-standardized ischemic heart disease DALY rates in Western Sub-Saharan Africa and globally, 1990–2023. DALY rates are expressed per 100,000 population and were obtained from the Global Burden of Disease 2023 study.

**Figure 5.**
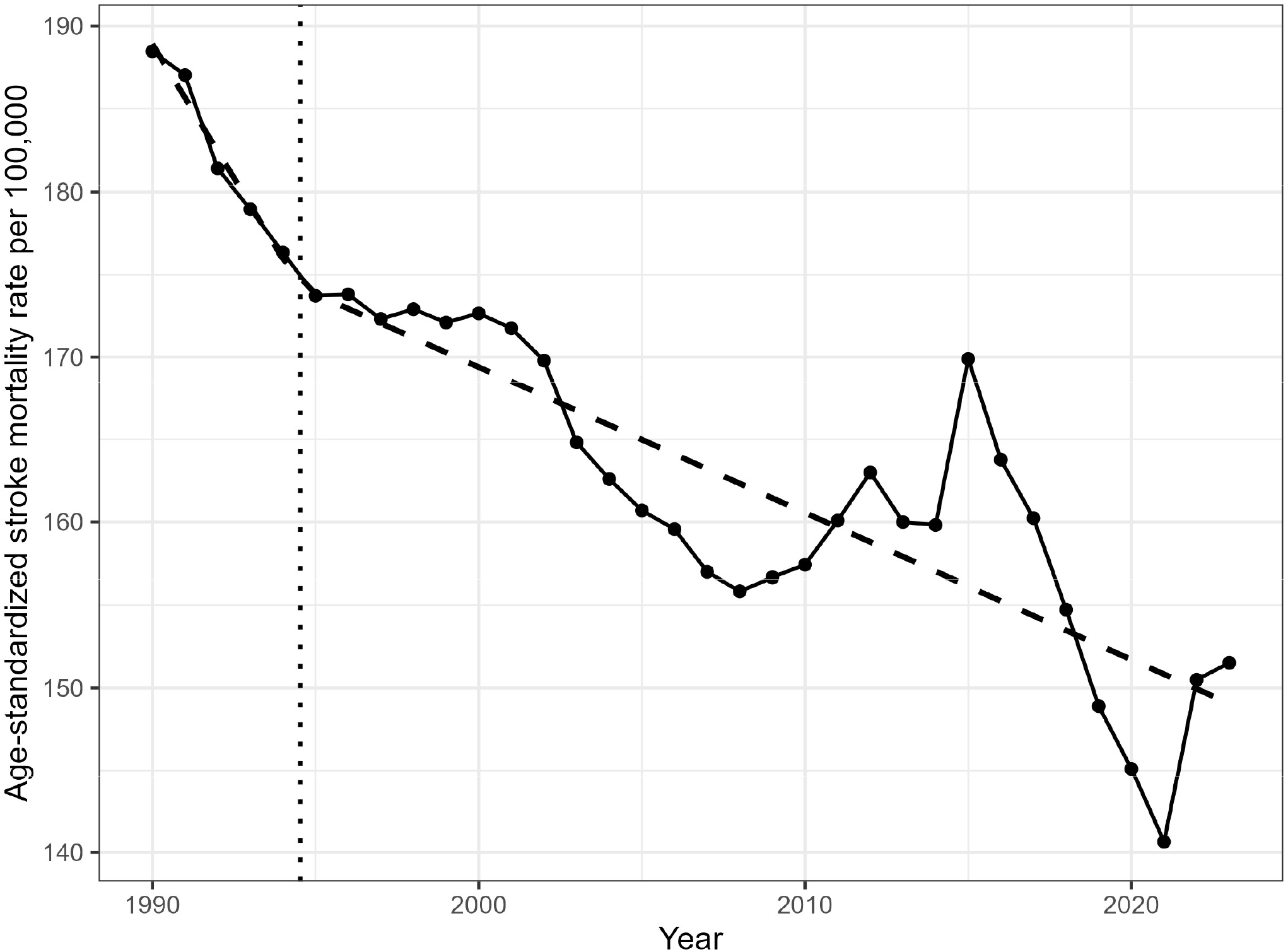
Segmented regression analysis of stroke mortality in Western Sub-Saharan Africa, 1990–2023. The vertical dotted line indicates the estimated breakpoint in 1994.5, after which the rate of decline in mortality slowed substantially.

**Figure 6.**
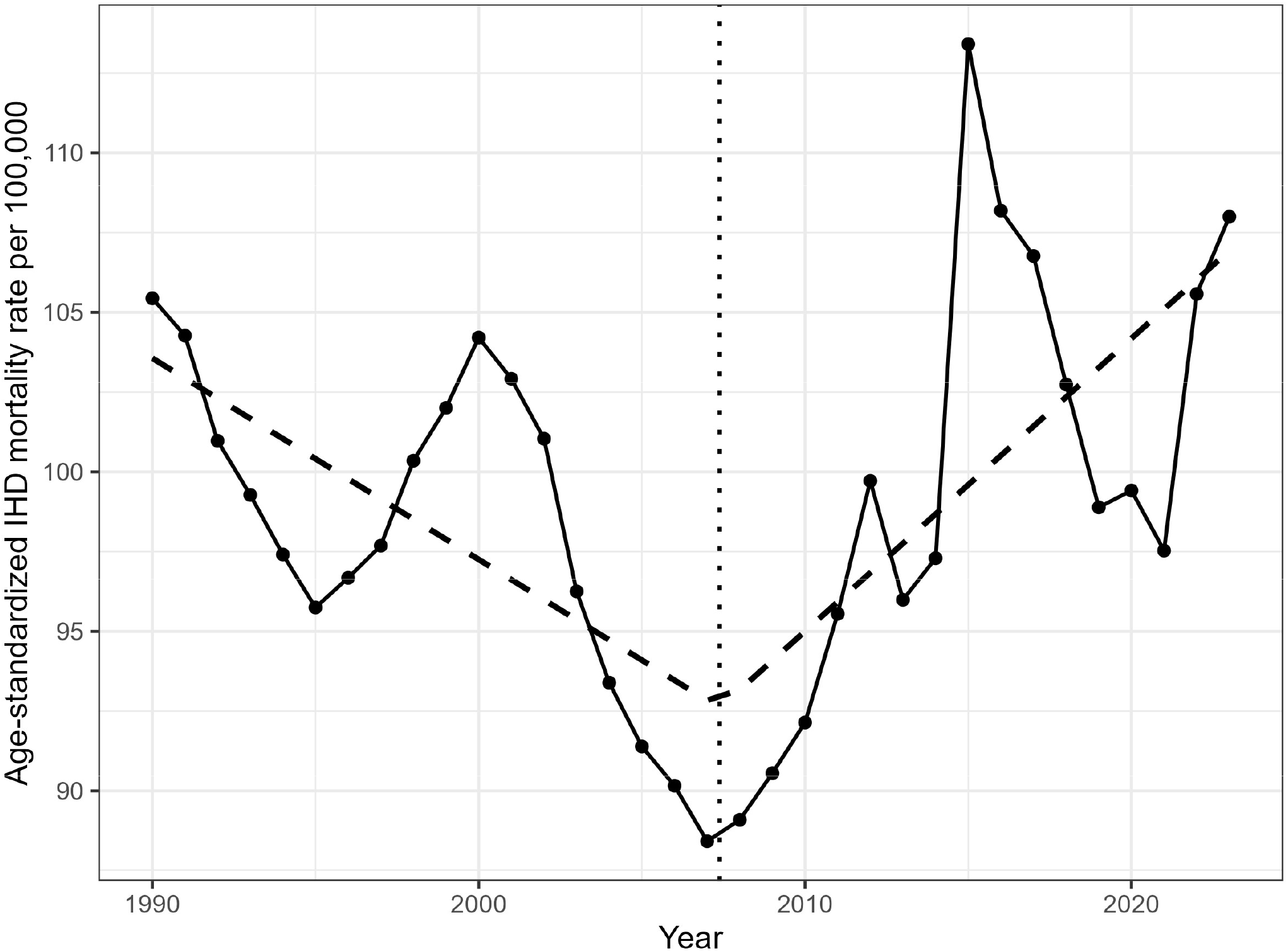
Segmented regression analysis of ischemic heart disease mortality in Western Sub-Saharan Africa, 1990–2023. The vertical dotted line indicates the estimated breakpoint in 2007.4, after which mortality trends reversed and began increasing.

### Risk-Factor Attribution in 2023

High systolic blood pressure was the leading attributable risk factor for both stroke and IHD in WSSA in 2023.

For stroke, the largest attributable burden was associated with high systolic blood pressure (mean rate 1889), followed by dietary risks (459), ambient particulate matter pollution (352), tobacco use (215), high fasting plasma glucose (192), and high body-mass index (187).

For IHD, high systolic blood pressure similarly represented the largest attributable burden (1122), followed by dietary risks (879), high body-mass index (300), ambient particulate matter pollution (277), tobacco use (214), and high fasting plasma glucose (144) (Table 4).

**Table 4.**
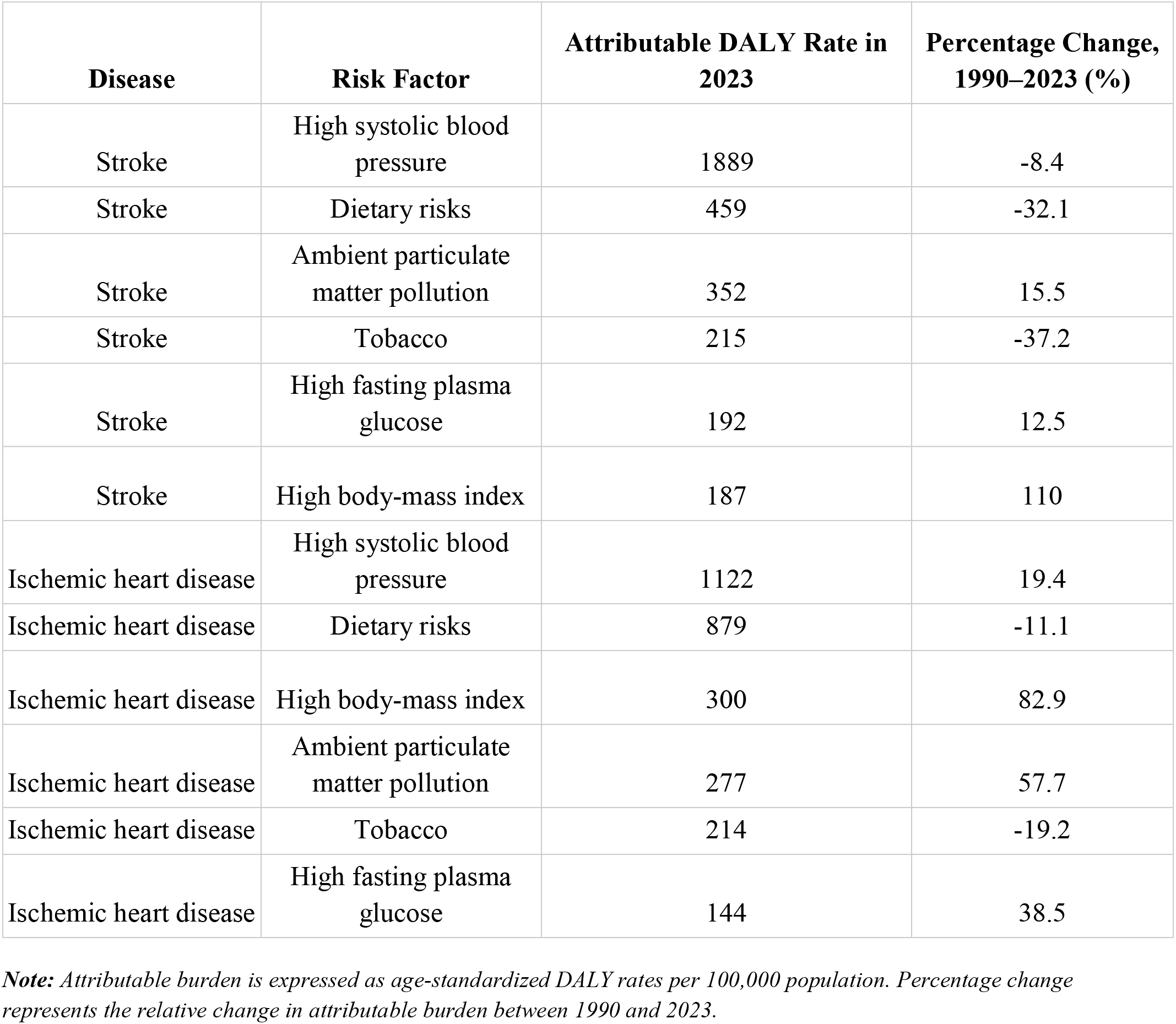
Leading Risk Factors and Attributable DALY Burden for Stroke and Ischemic Heart Disease in Western Sub-Saharan Africa, 2023, and Percentage Change Since 1990.

| Disease | Risk Factor | Attributable DALY Rate in 2023 | Percentage Change, 1990–2023 (%) |
| --- | --- | --- | --- |
| Stroke | High systolic blood pressure | 1889 | -8.4 |
| Stroke | Dietary risks | 459 | -32.1 |
| Stroke | Ambient particulate matter pollution | 352 | 15.5 |
| Stroke | Tobacco | 215 | -37.2 |
| Stroke | High fasting plasma glucose | 192 | 12.5 |
| Stroke | High body-mass index | 187 | 110 |
| Ischemic heart disease | High systolic blood pressure | 1122 | 19.4 |
| Ischemic heart disease | Dietary risks | 879 | -11.1 |
| Ischemic heart disease | High body-mass index | 300 | 82.9 |
| Ischemic heart disease | Ambient particulate matter pollution | 277 | 57.7 |
| Ischemic heart disease | Tobacco | 214 | -19.2 |
| Ischemic heart disease | High fasting plasma glucose | 144 | 38.5 |

**Note:** Attributable burden is expressed as age-standardized DALY rates per 100,000 population. Percentage change represents the relative change in attributable burden between 1990 and 2023.

Across both diseases, high systolic blood pressure contributed substantially more burden than any other individual risk factor.

### Changes in Risk-Factor Burden, 1990–2023

Substantial changes in cardiovascular risk-factor burden were observed over the study period.

For IHD, the largest increases were observed for high body-mass index (+82.9%), ambient particulate matter pollution (+57.7%), high fasting plasma glucose (+38.5%), and high systolic blood pressure (+19.4%). In contrast, attributable burdens related to dietary risks (−11.1%) and tobacco use (−19.2%) declined.

For stroke, high body-mass index exhibited the largest increase (+110.0%), followed by ambient particulate matter pollution (+15.5%) and high fasting plasma glucose (+12.5%). Conversely, attributable burdens associated with high systolic blood pressure (−8.4%), dietary risks (−32.1%), and tobacco use (−37.2%) decreased between 1990 and 2023 (Table 4).

These findings indicate increasing contributions from cardiometabolic and environmental risk factors despite reductions in several traditional cardiovascular risk factors (Figure 7).

**Figure 7.**
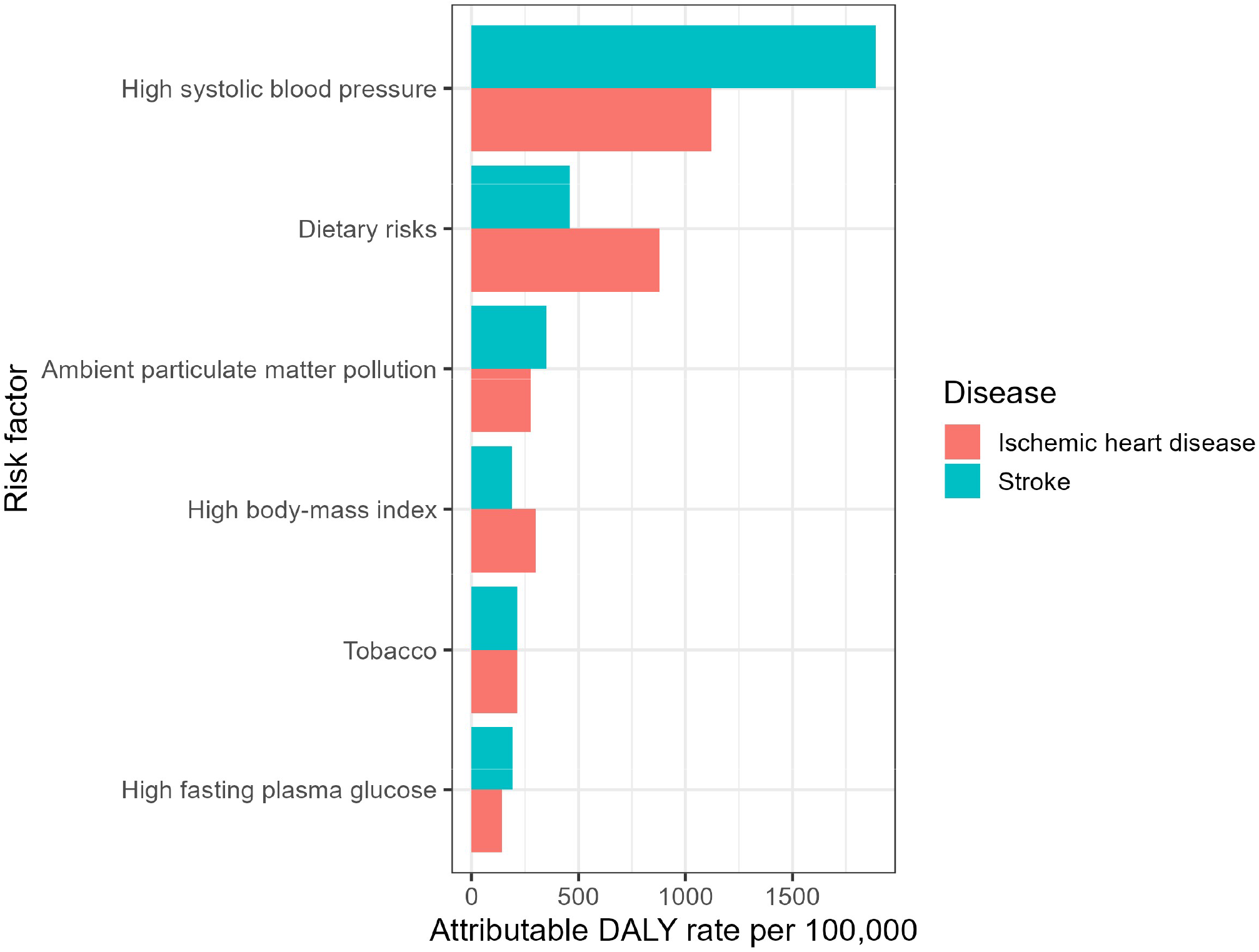
Major risk factor–attributable DALY rates for stroke and ischemic heart disease in Western Sub-Saharan Africa in 2023. DALY rates are expressed per 100,000 population. High systolic blood pressure was the leading attributable risk factor for both conditions.

### Socio-demographic Development and Age-Standardized Cardiovascular Mortality

Between 1990 and 2023, SDI increased substantially in WSSA, rising from 0.271 to 0.460, representing a 69.5% increase. Globally, SDI increased from 0.532 to 0.680, corresponding to a 28.0% increase (Table 5A).

**Table 5A.**
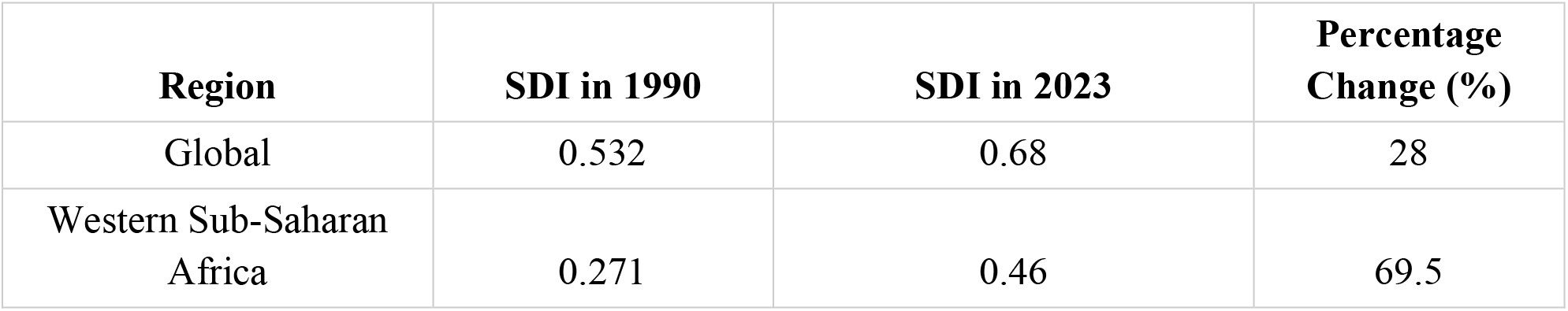
Changes in Socio-demographic Index (SDI), 1990–2023.

A strong inverse association was observed between SDI and age-standardized stroke mortality in WSSA (Table 5B; Pearson r = −0.866, 95% CI: −0.932 to −0.747, p < 0.001), indicating lower stroke mortality with increasing socio-demographic development.

**Table 5B.**
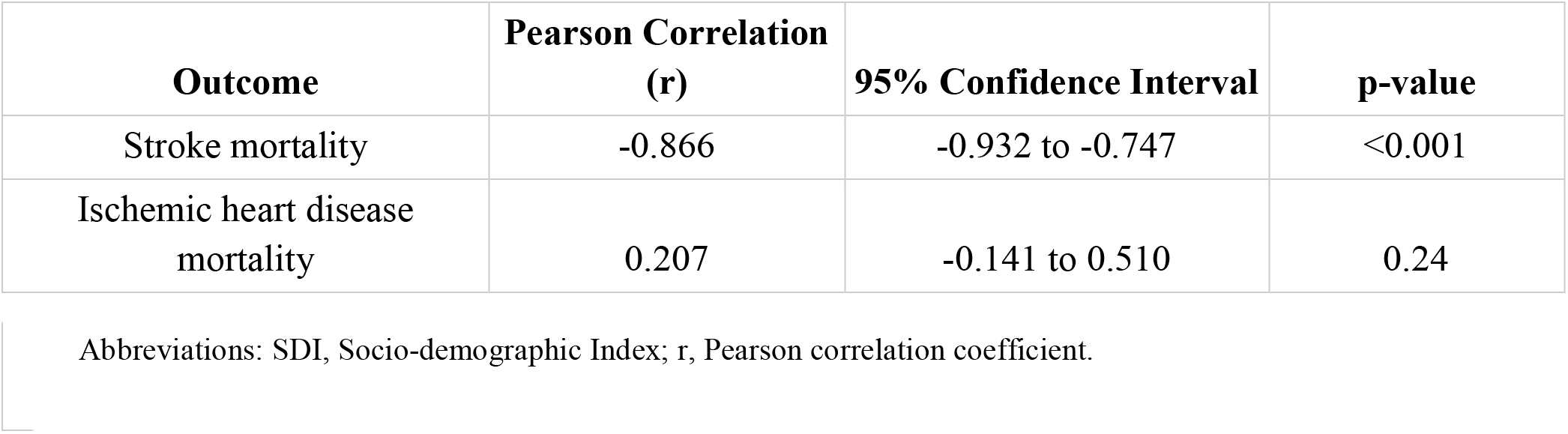
Correlation Between SDI and Age-Standardized Mortality Rates in Western Sub-Saharan Africa.

In contrast, no significant association was observed between SDI and age-standardized IHD mortality (Pearson r = 0.207, 95% CI: −0.141 to 0.510, p = 0.240). Despite substantial gains in socio-demographic development, improvements in age-standardized IHD mortality were not observed (Figure 8).

**Figure 8.**
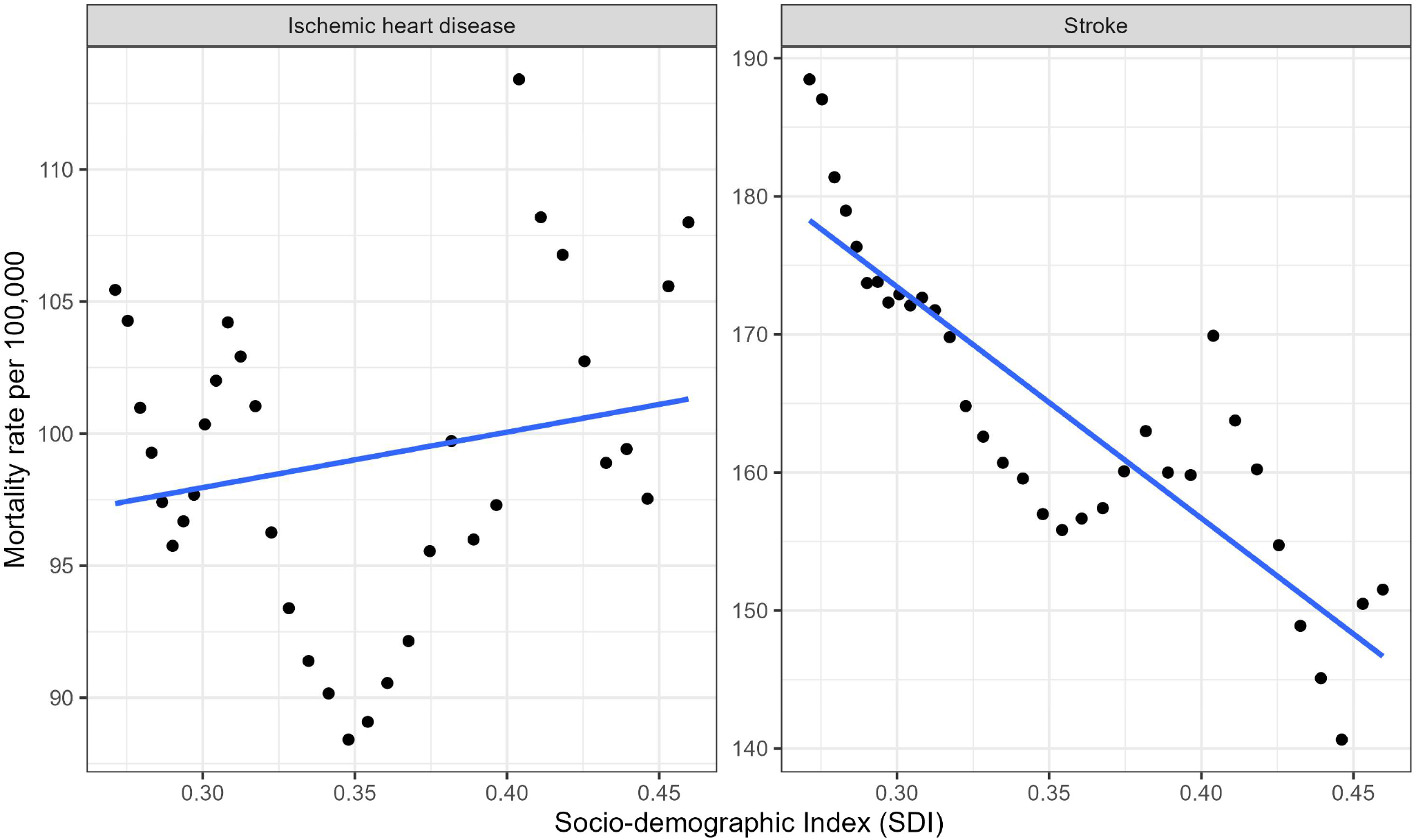
Association between socio-demographic index (SDI) and mortality rates in Western Sub-Saharan Africa, 1990–2023. Increasing SDI was strongly associated with declining stroke mortality but showed no significant association with ischemic heart disease mortality.

## 4. Discussion

This study reveals a striking divergence between Western Sub-Saharan Africa (WSSA) and the rest of the world in cardiovascular disease outcomes over the past three decades. Our analysis showed that while global age-standardized mortality rates for both stroke and ischemic heart disease (IHD) declined substantially between 1990 and 2023, progress in WSSA was markedly slower for stroke and entirely absent for IHD. The 51.7% global reduction in stroke mortality contrasts with only a 21.8% reduction in WSSA, and whereas global IHD mortality fell by 38.2%, WSSA experienced a 3.3% increase. Together, these findings indicate that WSSA is not merely progressing more slowly than the rest of the world but is following a distinctly different cardiovascular trajectory. This growing disparity is consistent with previous reports identifying Sub-Saharan Africa as the only world region where age-standardized cardiovascular mortality has failed to decline substantially. ⁴˒¹¹

The slower pace of stroke mortality decline in WSSA observed in our study is consistent with accumulating evidence that stroke burden in SSA is among the highest globally. Kim and Johnston demonstrated that stroke mortality rates exceeded IHD mortality rates in 74 of 192 countries, with lower national income strongly associated with higher relative stroke burden ^11^.

Feigin and colleagues reported that age-standardized stroke-related mortality was 3.6 times higher in low-income countries than in high-income countries in 2019 ^12^. Importantly, our segmented regression analysis in the present study identified a breakpoint around 1995, after which the rate of stroke mortality decline in WSSA decelerated substantially from β = −3.233 to β = −0.885 per year. This attenuation of progress may reflect the epidemiological transition in the region’s initial gains from early public health efforts and improvements in acute stroke care were not sustained as the underlying risk factor burden continued to grow ^13, 14^.

Several factors may explain the slowing of stroke mortality reductions observed in our analysis. Akinyemi and colleagues highlighted that Africa has up to 2-3-fold greater rates of stroke incidence and higher stroke prevalence than Western Europe and the USA, with hypertension as the dominant modifiable risk factor ^14^. The pooled prevalence of hypertension in SSA is approximately 27-30%, yet awareness, treatment, and control remain critically low, with fewer than one in 10 individuals achieving target blood pressure levels ^15–17^. Our finding that stroke mortality continued to decline at a much slower rate after 1995 is consistent with the persistent challenge of inadequate hypertension control across the region. Without effective blood pressure management at the population level, the stroke burden is likely to persist. Additionally, a higher proportion of hemorrhagic stroke in African populations constituting up to 35% of all strokes compared with 10-20% in high-income regions contributes to higher case fatality and limited responsiveness to conventional prevention strategies ^14,18^.

Perhaps the most alarming finding of this study is the reversal of IHD mortality trends in WSSA. Segmental regression analysis identified a breakpoint around 2007, after which IHD mortality shifted from a declining trajectory (β = −0.631) to a significantly increasing one (β = +0.918). To our knowledge, this temporal reversal has not been clearly demonstrated previously for WSSA using long-term GBD trend analysis. This finding is particularly concerning because it suggests that WSSA is beginning to experience a dual cardiovascular epidemic persistent stroke burden with a rising IHD burden superimposed on continued challenges from communicable diseases.

Our findings suggest that WSSA is entering a new phase of cardiovascular epidemiological transition characterized by a persistent stroke burden alongside an emerging IHD epidemic. The reversal in IHD mortality observed after 2007, despite continued declines globally, indicates that this transition is already underway rather than merely projected. This interpretation is consistent with predictions that the IHD burden in Africa would increase as epidemiological transition progresses. Onen projected that IHD mortality rates in SSA would rise by 70% in men and 74% in women by 2030 ^19^. Taha and colleagues described this phenomenon as an “overnight epidemiological transition,” noting that IHD ranked as the leading cause of death for men and the second leading cause for women over 60 years in the African region ^5^. Moran and colleagues, using GBD 2010 data, showed that while age-standardized IHD mortality declined in most world regions, it increased in several low-and middle-income regions ^20^. Xia and colleagues specifically identified Western Sub-Saharan Africa as one of the regions where IHD incidence increased between 1990 and 2019, associated with rapid socioeconomic transition ^6^.

The rising IHD mortality in WSSA may reflect the accelerating burden of atherogenic risk factors. Historically, IHD was relatively uncommon in SSA, partly attributed to favorable lipid profiles, low total cholesterol and high HDL cholesterol among African populations ^21, 22^. However, urbanization and westernization of lifestyles are altering this protective profile. Vorster demonstrated that increasing total fat and animal protein intake among affluent urban South Africans was associated with rising body mass index and total serum cholesterol, predicting future increases in IHD risk ^21^. The present study’s finding that high body-mass index attributable DALY burden increased by 82.9% for IHD between 1990 and 2023 provides direct evidence supporting the growing contribution of adiposity to the region’s evolving IHD burden.

Our risk factor analysis revealed a critical shift in the cardiovascular risk profile of WSSA. High systolic blood pressure remained the dominant attributable risk factor for both stroke and IHD in 2023, consistent with decades of literature establishing hypertension as the leading cardiovascular risk factor in Africa ^23–25^. However, the temporal changes in attributable burden were divergent: traditional risk factors such as tobacco use and dietary risks showed declining attributable burden, while cardiometabolic and environmental risk factors showed substantial increases.

The most dramatic increase was in high body-mass index, which rose by 110.0% for stroke and 82.9% for IHD. This aligns with the GBD 2019 global analysis identifying high body-mass index as the fastest-growing risk factor for stroke worldwide ^12^. The marked increase in BMI-attributable burden observed in our study suggests that obesity is becoming an increasingly important driver of cardiovascular disease in WSSA particularly as the region undergoes rapid nutritional and lifestyle transitions. The obesity epidemic in SSA is driven by rapid urbanization, nutrition transition characterized by increasing consumption of energy-dense processed foods, and declining physical activity ^26–28^. Ziraba and colleagues documented that urban overweight and obesity prevalence in Africa increased by nearly 35% between the 1990s and 2000s, with the increase most pronounced among the poorest and least-educated women ^27^. Peer and colleagues noted that obesity prevalence in SSA varies considerably but is rising rapidly, with prevention and control remaining disorganized and insufficient ^29^.

Ambient particulate matter pollution attributable burden also increased substantially by 57.7% for IHD and 15.5% for stroke. Air pollution in Africa is an increasingly recognized but understudied cardiovascular risk factor. Katoto and colleagues found that ambient air pollution levels in SSA cities were 10-20 fold higher than WHO standards, with very limited monitoring infrastructure ^30^. Adeoye and colleagues highlighted the critical need for standardized research on air pollution and cardiovascular health in Africa, noting the extremely limited number of studies ^31^. In WSSA specifically, rapid urbanization, reliance on solid biomass fuels, expanding vehicle fleets composed of imported used vehicles, and open waste burning contribute to rising exposure.

High fasting plasma glucose attributable burden increased by 38.5% for IHD and 12.5% for stroke. The rising diabetes burden in SSA parallels the obesity epidemic and is exacerbated by late diagnosis, poor management, and limited access to treatment ^31, 33^. These metabolic risk factors as components of the metabolic syndrome operate synergistically with hypertension to accelerate atherosclerosis and increase cardiovascular event risk ^34^.

In contrast, tobacco use attributable burden declined (−37.2% for stroke, −19.2% for IHD). While this is encouraging, it should be interpreted cautiously. Tobacco use remains a significant risk factor, and the declining attributable burden may reflect differences in the relative contribution of other rapidly rising risk factors rather than absolute reductions in tobacco-related harm. Furthermore, tobacco industry targeting of African markets has been documented, and consumption may still increase as economies develop ^35, 36^.

The divergent associations between SDI and disease-specific mortality in WSSA provide important insights into the region’s epidemiological transition. While SDI showed a strong inverse correlation with stroke mortality (r = −0.866, p < 0.001), no significant association was observed with IHD mortality (r = 0.207, p = 0.240). This pattern is consistent with the classical model of epidemiological transition, where stroke (associated with hypertension and its complications) initially declines with socioeconomic development, while IHD may increase as populations adopt westernized diets and sedentary lifestyles ^7,28,37^.

Our findings suggest that socioeconomic development alone is insufficient to improve ischemic heart disease outcomes in WSSA. Despite substantial gains in SDI over the past three decades, these improvements were not accompanied by a corresponding decline in IHD mortality, indicating that broader socioeconomic progress may not translate into cardiovascular benefit in the absence of parallel investments in prevention, diagnosis, and treatment. This interpretation is consistent with findings from the PURE study, which demonstrated that individuals in low-income settings often have more favorable cardiovascular risk profiles yet experience poorer outcomes because of limited access to healthcare services and evidence-based management ^38^. Similarly, Okorigba and colleagues recently reported that persistently high IHD mortality in Sub-Saharan Africa closely parallels limited access to interventional cardiology services and preventive cardiovascular programs ^39^.

The 69.5% increase in SDI in WSSA between 1990 and 2023 indicates substantial socioeconomic advancement, yet this development has not translated into improved IHD outcomes. This paradox may be explained by the fact that rapid socio-economic development in the region has outpaced the capacity of health systems to respond to the emerging cardiovascular disease burden. Health systems in SSA remain predominantly oriented toward addressing communicable diseases, with limited infrastructure and expertise for cardiovascular prevention and treatment ^8, 29, 40^. Dzudié and colleagues noted that limited availability of clinical expertise, diagnostic facilities, and access to optimal medical therapy, combined with lack of universal health coverage, constitute major challenges for chronic coronary syndrome care in SSA ^41^.

The differing trajectory of stroke and IHD mortality in WSSA highlights that these two conditions, though sharing risk factors, represent distinct epidemiological challenges in the SSA context. Stroke has been the predominant cardiovascular killer in SSA for decades, with more deaths from stroke (409,840) than IHD (258,939) reported in 2013 ^3^. Kim and Johnston demonstrated that stroke burden was disproportionately higher in lower-income countries, while IHD burden was higher in the Middle East, North America, and much of Europe ^11^.

The stronger association between SDI and stroke mortality may reflect the greater potential for population-level blood pressure control to reduce stroke incidence and mortality. In the Seychelles one of the few SSA countries with comprehensive vital registration, Stringhini and colleagues demonstrated that both stroke and myocardial infarction mortality declined substantially between 1989 and 2010, with declines of 44%/39% for stroke and 50%/53% for MI ^42^. This success was attributed to comprehensive vital registration and systematic cardiovascular disease prevention. The Seychelles example suggests that SSA countries can achieve meaningful cardiovascular mortality reductions when functional health systems and surveillance are in place.

In contrast, IHD prevention and management require more complex interventions, including management of dyslipidemia, advanced cardiac imaging, interventional cardiology, and sophisticated pharmacotherapy services that are scarce in most WSSA countries ^41, 43^. Yusuf and colleagues reported that use of secondary prevention medications for cardiovascular disease was lowest in low-income countries, where 80.2% of patients received no medications at all ^44^. This severe treatment gap may partially explain the rising IHD mortality despite increasing SDI.

### Implications for policy and practice

The findings of this study carry several important policy implications. First, hypertension control must remain the cornerstone of cardiovascular disease prevention in WSSA. The data confirm that high systolic blood pressure is the leading attributable risk factor for both stroke and IHD, yet control rates across SSA remain catastrophically low ^16, 17,45^. Etyang and colleagues have called for a dual agenda of implementation science scaling proven interventions such as task-sharing with community health workers, single pill combination therapies, and mHealth platforms and discovery research to understand region-specific risk factors and disease patterns ^17^. Community-based screening programs, affordable antihypertensive medications, and integration of hypertension management into existing primary healthcare and HIV programs represent cost-effective strategies ^35, 36, 46^.

Second, the rapidly rising burden of obesity, diabetes, and metabolic syndrome in WSSA demands urgent action. Population-level interventions targeting dietary salt reduction, food labeling, taxation of sugar-sweetened beverages, and promotion of physical activity have been recommended by the WHO Global Action Plan for NCDs and have proven cost-effective in other settings ^35, 36^. However, only South Africa in the SSA region has a national program to reduce dietary salt intake ^35,36^. The unchecked expansion of Western fast-food chains and processed food marketing on the continent further accelerates the nutrition transition ^47^.

Third, the rising burden of ambient particulate matter pollution attributable to cardiovascular disease necessitates environmental health interventions. Addressing household air pollution through clean cooking energy transitions, improving vehicle emission standards, and strengthening air quality monitoring networks are essential ^31,48, 49^.

Fourth, health system strengthening is imperative. The gap between rising cardiovascular disease burden and health system capacity in SSA is widening. Doku and colleagues demonstrated that in Ghana, fewer than 10% of hypertensive patients had controlled blood pressure, with suboptimal health worker knowledge, lack of standardized CVD management protocols, and limited diagnostic equipment ^43^. The Pan-African Society of Cardiology and the World Heart Federation have advocated for task-shifting, training of cardiovascular healthcare workers, and development of context-appropriate treatment guidelines ^50, 51^. Recent WHO HEARTS initiatives in SSA have shown promising progress and represent scalable models for cardiovascular risk management ^17^.

Finally, improved vital registration and disease surveillance are essential for monitoring trends and evaluating interventions. The reliance on modeled GBD estimates rather than directly enumerated data remains a limitation for all analyses of cardiovascular disease burden in SSA ^3,10,52^. Investments in civil registration and vital statistics systems, along with population-based epidemiological studies, are critical for informing data-driven health policy ^8,53^.

## 5. Strengths and Limitations

This study benefits from 33 years of data spanning 1990 to 2023, providing a comprehensive longitudinal perspective on cardiovascular disease trends in WSSA. The use of GBD 2023 data ensures consistency and comparability with global estimates. Multiple analytical approaches linear regression, segmented regression, correlation analysis, and risk factor attribution provide complementary insights into the dynamics of cardiovascular disease in the region. The direct comparison between WSSA and global trends highlights disparities that regional analyses alone might not reveal.

However, several limitations should be acknowledged. As an ecological analysis, this study examines population-level associations and cannot infer individual-level causation. The GBD estimates used rely on modeled data rather than directly observed clinical records, and their accuracy depends on the quality and availability of underlying data sources which vary substantially across WSSA countries ^3, 10^. Missing or sparse vital registration data in several WSSA countries means that GBD estimates incorporate substantial uncertainty, particularly for earlier years of the study period ^52^.

The analysis does not distinguish between stroke subtypes (ischemic vs. hemorrhagic), which have different risk factor profiles and prognoses, and the proportion of hemorrhagic stroke may be higher in WSSA than globally ^14, 18^.

Individual-level clinical variables including treatment patterns, access to care, medication adherence, and healthcare-seeking behavior cannot be captured in ecological analyses. The risk factor attribution analysis examines DALYs rather than direct causal pathways, and the contribution of risk factors may be interrelated.

Finally, the SDI-mortality correlation analysis cannot establish causality and may be confounded by other socioeconomic or environmental variables not captured in the SDI composite.

## 6. Conclusion

This study reveals a clear divergence between Western Sub-Saharan Africa and global cardiovascular progress over 1990–2023. While global stroke and ischemic heart disease mortality declined substantially, WSSA saw only modest stroke gains and a reversal in IHD mortality after 2007, despite a 69.5% rise in socio-demographic development. High systolic blood pressure remained the dominant risk factor for both conditions, while obesity, air pollution, and metabolic risk burden rose sharply, suggesting the region’s epidemiological transition is outpacing its health system capacity. The divergent SDI associations for stroke and IHD indicate that socioeconomic development alone will not resolve this gap; the mechanisms driving each disease differ enough that a single prevention strategy is unlikely to address both. Regional policymakers and health systems must act now to scale hypertension control, address rising cardiometabolic and environmental risk factors, and expand cardiovascular care capacity, before WSSA’s dual burden of persistent stroke mortality and rising IHD mortality becomes further entrenched.

## Acknowledgments

The authors acknowledge the University of Aberdeen for its support of scholarly publishing through institutional agreements that facilitate open-access publication. The authors also acknowledge the Institute for Health Metrics and Evaluation (IHME) and collaborators of the Global Burden of Disease 2023 Study for making the data used in this analysis publicly available.

## Author Contributions

Prince Ankrah-Twumasi: Conceptualization; Methodology; Formal analysis; Visualization; Writing – original draft.

Jeffrey Jerry Vladimir Ofori: Formal analysis; Writing – original draft; Writing – review & editing.

Prince Pekyi-Boateng: Conceptualization; Methodology; Formal analysis; Writing – original draft.

Yaw Twerefour: Formal analysis; Methodology; Validation; Writing – review & editing. Dorcas Sackey: Conceptualization; Methodology; Writing – review & editing.

All authors contributed to the revision of the manuscript, approved the final version to be published, and agree to be accountable for all aspects of the work.

## Data Availability Statement

All data used in this study were obtained from the Global Burden of Disease (GBD) 2023 Study and are publicly available through the Institute for Health Metrics and Evaluation (IHME) data repository. The datasets analyzed during the current study are available from the corresponding sources and can be accessed without restriction.

## Ethics Statement

Ethics approval was not required for this study because it involved secondary analysis of publicly available, de-identified data from the Global Burden of Disease 2023 Study.

---

**Table S1.** Western Sub-Saharan African Countries Included in the Analysis.

| Countries |
| --- |
| Benin |
| Burkina Faso |
| Cabo Verde |
| Côte d'Ivoire |
| Gambia |
| Ghana |
| Guinea |
| Guinea-Bissau |
| Liberia |
| Mali |
| Mauritania |
| Niger |
| Nigeria |
| Senegal |
| Sierra Leone |
| Togo |

Note: Country classification was based on the Global Burden of Disease (GBD) 2023 regional grouping for Western Sub-Saharan Africa.

